# Impact of COVID-19 Lockdown Policy on Homicide, Suicide, and Motor Vehicle Deaths in Peru

**DOI:** 10.1101/2020.07.11.20150193

**Authors:** Renzo JC Calderon-Anyosa, Jay S Kaufman

**Affiliations:** Department of Epidemiology, Biostatistics and Occupational Health, McGill University, Montreal, Canada

## Abstract

**Introduction:** Although lockdown measures to stop COVID-19 have direct effects on disease transmission, their impact on violent and accidental deaths remains unknown. Our study aims to assess the early impact of COVID-19 lockdown on violent and accidental deaths in Peru.

**Methods:** Based on data from the Peruvian National Death Information System, an interrupted time series analysis was performed to assess the immediate impact and change in the trend of COVID-19 lockdown on external causes of death including homicide, suicide, and traffic accidents. The analysis was stratified by sex and the time unit was every 15 days.

**Results:** All forms of deaths examined presented a sudden drop after the lockdown. The biggest drop was in deaths related to traffic accidents, with a reduction of 12.66 deaths per million men per month (95% CI: −15.56, −9.76) and 3.64 deaths per million women per month (95% CI:-5.25, −2.03). Homicide and suicide presented similar level drop in women, while the homicide reduction was twice the size of the suicide reduction in men. The slope in suicide in men during the lock-down period increased by 3.62 deaths per million men per year (95% CI:0.06, 7.18). No other change in slope was detected.

**Conclusions:** Violent and accidental deaths presented a sudden drop after the lockdown was implemented and an increase in suicide in men was observed. Falls in mobility have a natural impact on traffic accidents, however, the patterns for suicide and homicide are less intuitive and reveal important characteristics of these events, although we expect all of these changes to be transient.

## Introduction

COVID-19 has had a serious impact on population health worldwide ^1^, not only as a direct consequence of the infection but also due to the measures taken to reduce its transmission. These unprecedented changes in the lifestyles of millions have also impacted mental health, society and economy in various ways ^2,3^.

The main strategies to reduce COVID-19 transmission are social distancing and isolation measures. Policies range from advising individuals to keep apart a physical distance (i.e. 2 meters) in public spaces all the way to generalized lockdowns ^4^, all to reduce the pace of transmission and to prevent health services from being overwhelmed ^5^.

By the middle of June 2020, Latin-America had become a focus of COVID-19 infection. To slow the spread, most of the countries in the region have taken severe lockdown measures ^6^. Peru was one of the countries with the earliest and strictest national lockdown measures in Latin-America and has won international recognition for its pandemic response, starting restrictions right after the first confirmed case in mid-March 2020 and lasting for over 100 days until the end of June in most of the national territory ^6–9^.

Although measures such as national lockdowns are expected to have a direct impact on transmission and subsequent mortality due to COVID-19 infection^10–12^, their interruption of all daily activities suggest that they may also impact other aspects of health and causes of death ^13–15^. Radical changes in the daily activities of individuals, self-isolation, financial uncertainty, job losses, and reduced incomes have the potential to influence mortality due to violent crimes, suicides, domestic violence, and other external causes of death ^16–19^.

Early reports have indicated a substantial drop in violent crime rates across the world, with an overall drop in crime close 50% in some cities following some of the most restrictive measures ^20,21^. At the same time, reports of domestic violence have increased since social distancing measures came into effect, as victims are forced to be isolated with their abusers^22,23^. The UN has estimated that domestic violence has increased by over 30% in some countries since lockdown with a surge in the need for shelters^24,25^.

Lockdowns have also been accompanied by travel bans and a reduction in mobility, leading to a decrease in the use of motor vehicles use^26^ with a consequential drop in traffic accidents and resultant emergency visits and deaths^27,28^. The mental health effects of COVID-19 and of the accompanying economic crisis have also been profound, with suicide as a concern^16^. Previous pandemic scenarios have also shown a change in suicide trends including the 1918 influenza pandemic and 2003 SARS epidemic^29,30^.

Although by the end June 2020 most countries have relaxed their lockdown measures, their diverse consequences are still unclear and are just now beginning to be studied empirically. Our study aims to assess the early impact of the COVID-19 national lockdown on homicide, suicide, and traffic accident deaths in the Peruvian setting.

## Methods

### Study population

We used data from the Peruvian National Death Information System (SINADEF)^31^ which collects daily death certificates nationwide with available data since 2017. SINADEF has improved the quality of data registration in the recent years, managing to improve its coverage and represents close to 80% of all deaths in the national territory^32^. For this study, we have included nationwide information on deceased adults (18 years old or older) from January 1^st^, 2017 to June 28^th^, 2020.

On March 16, 2020, the Peruvian government decreed a state of sanitary emergency, suspending economic, academic, and recreational activities across the entire country of 32 million people. Only essential activities including food supply, pharmacies, and banking remained active. Moreover, international borders were closed, military and police patrolled the streets, and a curfew was instituted from 8 p.m. to 5 a.m. Public transport capacity was also reduced by half and movement between regions within the country was banned. Although some of the components of the lockdown have changed throughout its implementation, the core aspects of the lockdown remained constant until the end of June 2020 ^7,8^.

### Measures

Because the lockdown was implemented in the middle of March and there is a relatively low count of daily deaths, we chose to aggregate the data in bins of 15 days each to have a uniform time unit throughout our study period.

Violent and accidental death information, sex, and age were taken directly from the SINADEF report. This report has an independent item for violent forms of deaths including homicide, suicide, traffic accidents, and other types of accidental deaths. This information is recorded by the health worker that certifies the death. The number of events was transformed into the rate per 1,000,000 population for better comparison based on the population report from the latest National Census^33^.

Because the COVID-19 pandemic imposes additional stress on health workers and the health system, the reporting, recording, and coding of deaths could be affected by this overload of work^34,35^. To estimate if violent deaths recording was affected by this scenario, we also estimated the proportion of violent deaths labeled as “unspecified” as a proportion of the total violent deaths. This fraction was assessed in the same way as the main outcomes to find any change in the trends after lockdown.

To have a measure of the degree of compliance with lockdown and an approximation to the use of motor vehicles we used descriptive data from the mobile-phone mobility data provided by Google Community Mobility Reports for public transit places^36^. This report presents the percent change in visits to transport places for each day compared to a baseline value for that day of the week.

### Statistical analysis

To assess the immediate impact and change in the trend of COVID-19 lockdown, we analyzed the violent death rates per population using an interrupted time series analysis^37^. A linear regression model was fitted to the violent deaths rates with a time variable (every 15 days), a variable to indicate post-lockdown, which was defined since March 16, 2020, and an interaction term between the post-lockdown indicator and the time variable, to evaluate a change in the slope of the outcome trend after lockdown. Stratified analysis was performed for women and men because of known differences in violent deaths by sex. Autocorrelation of the time series was assessed through a correlogram and seasonality through visual inspection of the plots. The analysis was conducted using R 3.6.1.

## Results

A total of 388,772 events were identified as adult deaths from January 1^st^, 2017 to June 28^th^, 2020. Violent and accidental deaths sum up to 14,526 including 6,700 traffic accidents, 2,849 homicides, 1,611 suicides, and 3,366 other forms of accidental deaths. No autocorrelation or seasonality was found in any time series because the entire follow-up period spanned only 3.5 months.

The time slope in the pre-lockdown period was positive for all types of violent deaths in both sex groups with the highest for male traffic accidents, with an extra 0.75 deaths per year per million men (95%CI: 0.51, 0.99) and the lowest in female homicides, with an extra 0.10 deaths per year per million women (95%CI: 0.04, 0.16). All forms of violent and accidental deaths presented a sudden drop after the implementation of the lockdown in both groups. The biggest difference in the post-lockdown period was in deaths related to traffic accidents, with a reduction of 12.66 deaths per million men per month (95% CI: −15.56, −9.76) and a reduction of 3.64 deaths per million women per month (95% CI:−5.25, −2.03) after the lockdown. Homicide and suicide presented a similar level drop in women with around 1 fewer death per million women per month, while the homicide reduction was twice the suicide reduction in men with 6 and 3 drops in deaths per million men per month respectively. Other forms of accidental deaths in women presented a reduction of 2 deaths per million women per month, being the second-highest drop in this group (Table 1). We detected an increase in the slope of suicides in men in the post-lockdown period with an extra 3.62 deaths per million men per year (95%CI: 0.06, 7.18) from the pre-lockdown period slope. No other change in the post-lockdown slope compared to the pre-lockdown period was found in the other types of violent or accidental death in women (Figure 1) or men (Figure 2). The post-lockdown follow-up is short and so there is low power to detect a slope change among these points^38^.

**Table 1.**
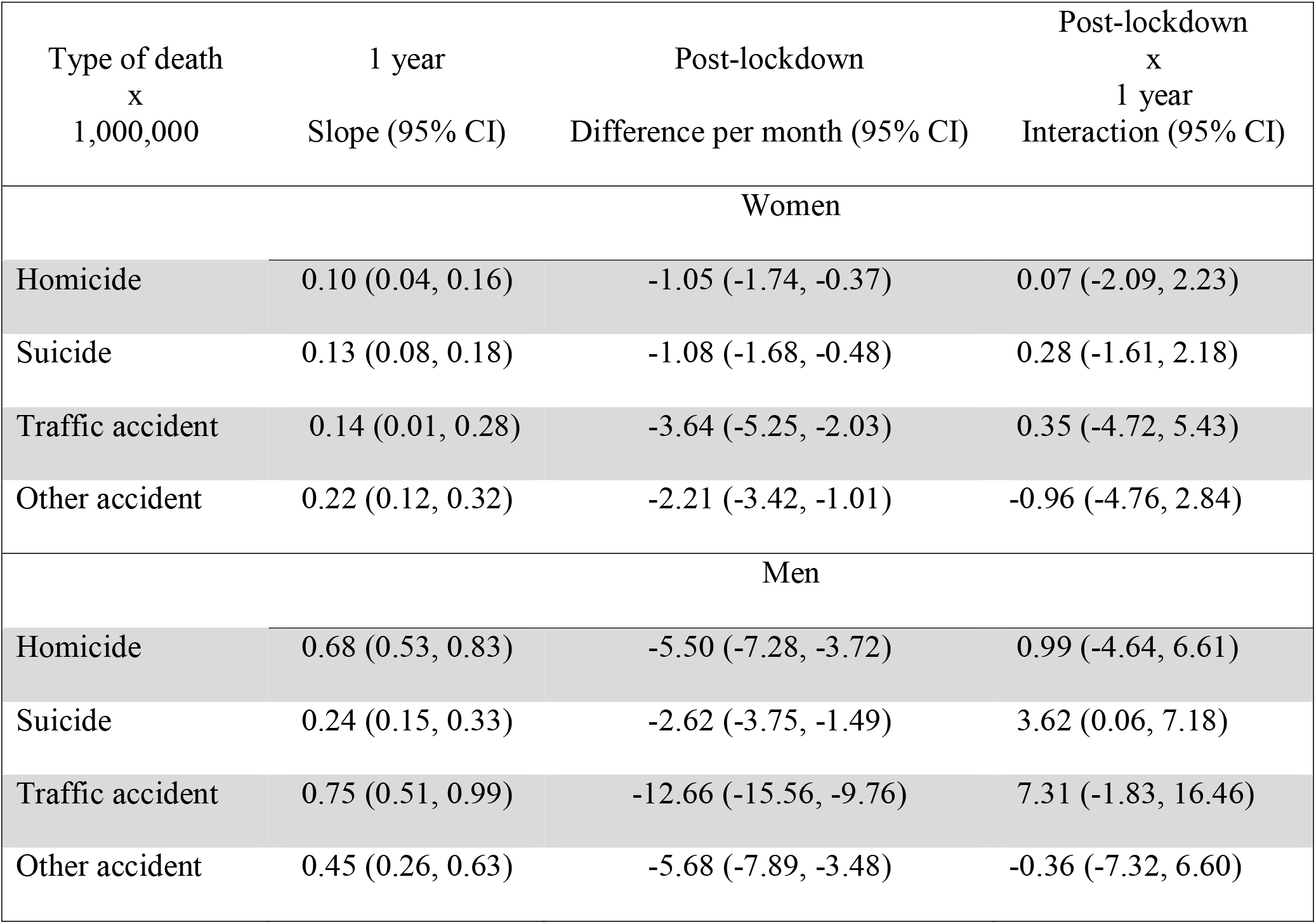
Interrupted time series coefficients by sex and type of violent or accidental death

**Figure 1.**
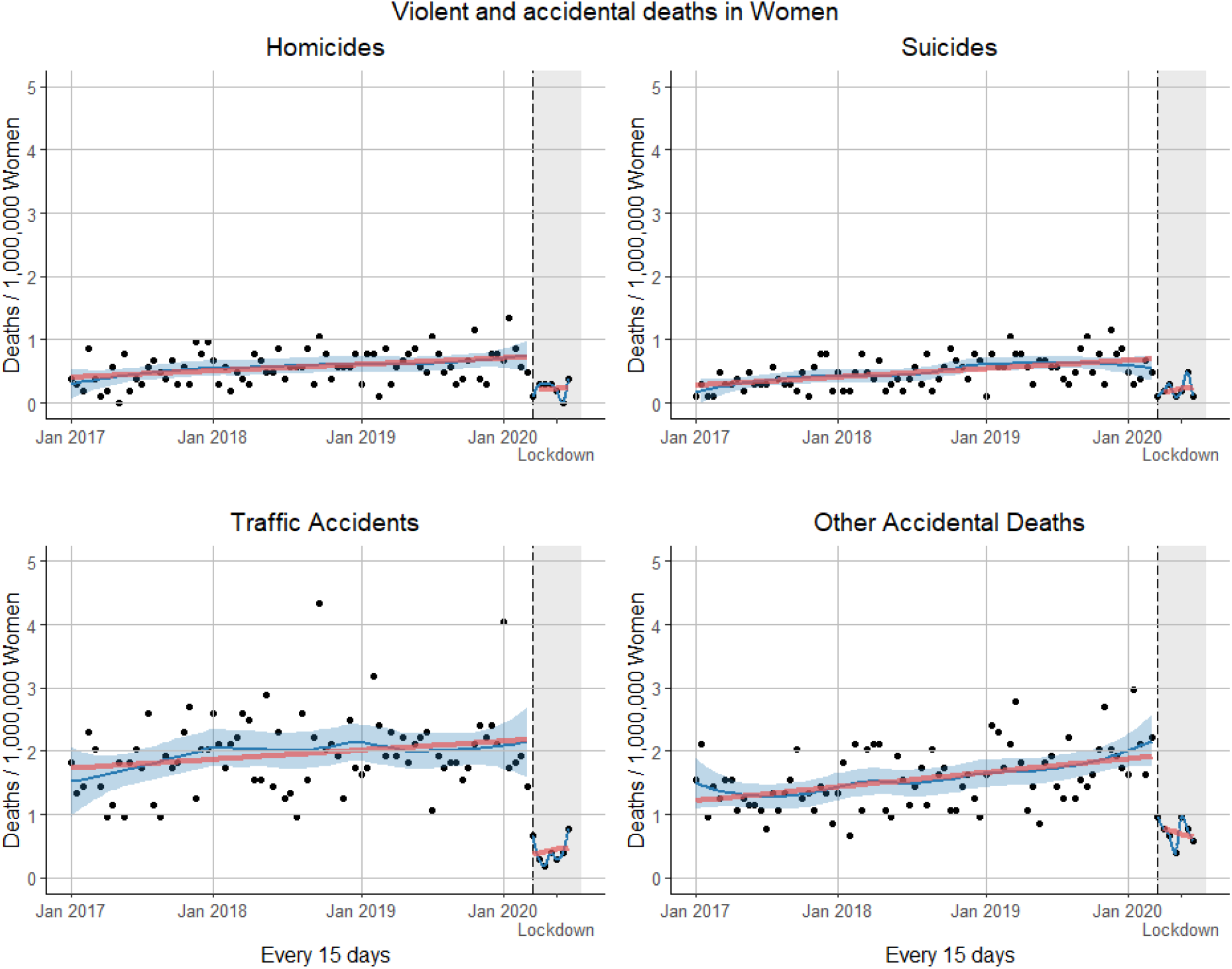
Interrupted time-series analysis violent deaths by type of death in Women. The vertical black dashed line corresponds to the beginning of the lockdown (March 16, 2020) and the grey shading to the lockdown period. The solid red lines correspond to the interrupted time-series linear regression model, the solid blue line corresponds to the LOESS smoother and the blue shading corresponds to a 0.6 smoothing span around the LOESS smoother line.

**Figure 2.**
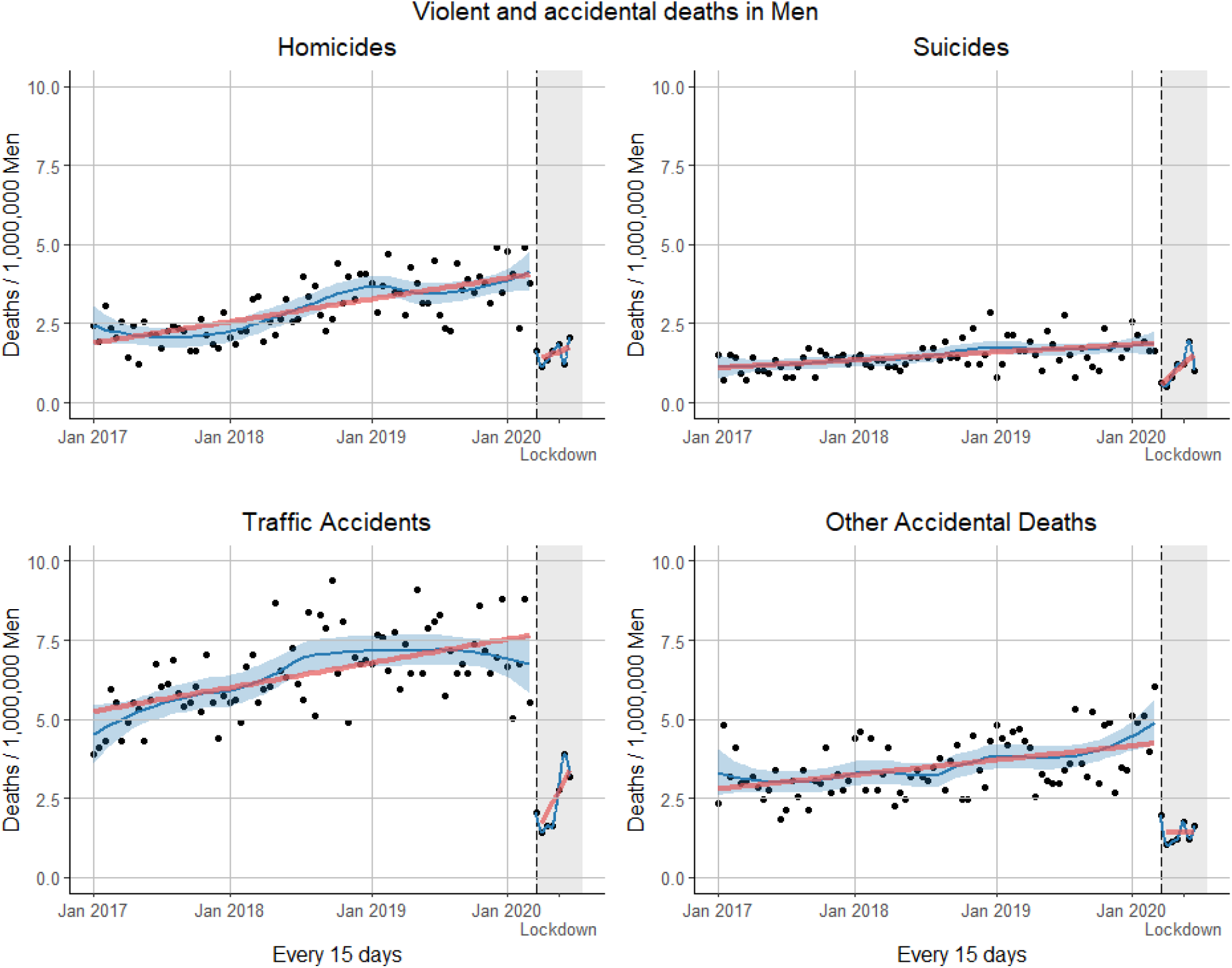
Interrupted time-series analysis violent deaths by type of death in Men. The vertical black dashed line corresponds to the beginning of the lockdown (March 16, 2020) and the grey shading to the lockdown period. The solid red lines correspond to the interrupted time-series linear regression model, the solid blue line corresponds to the LOESS smoother and the blue shading corresponds to a 0.6 smoothing span around the LOESS smoother line.

There was no change in the level or the trend of the unspecified violent death proportion (Figure 3A). The mobility data (Figure 3B) shows an early drop in mobility to transit stations right after the first confirmed case with a gradual reduction until the start of the lockdown. After lockdown, the mobility fell below −75% after the second day and held constant at around −80% for 40 days. After that period, mobility gradually recovered, with an increasing tendency through the end of the lockdown. The episodic drops shown close to −100% represent the strict curfew on Sundays and holidays.

**Figure 3.**
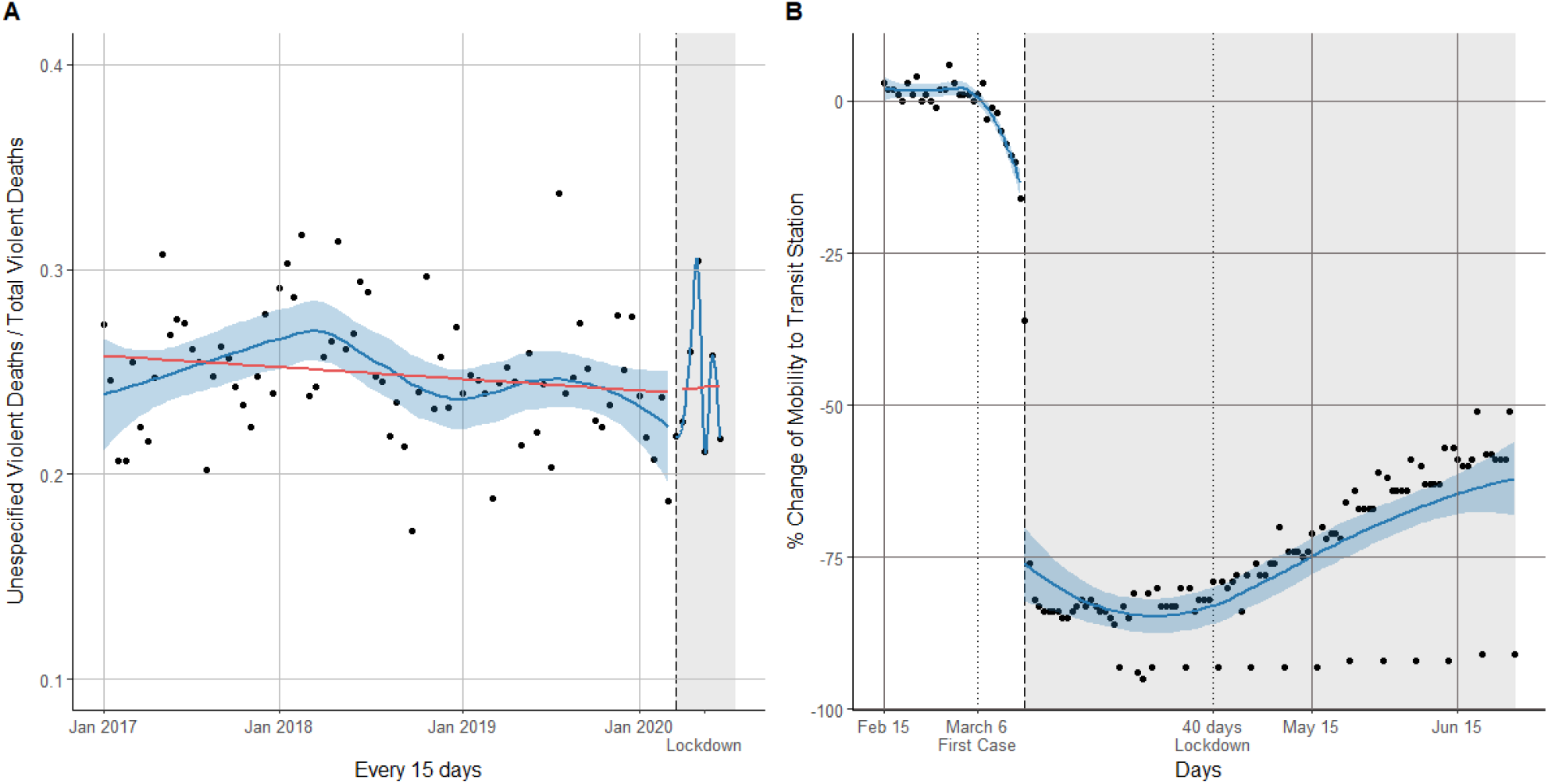
Panel A: Interrupted time-series analysis the propotion of violent deaths labeled as “unescpecified”. The solid red lines correspond to the interrupted time-series regression model. Panel B: Descriptive view of the percentage change in community mobility to transit stations before and after the lockdown period. The first vertical black dotted line corresponds to the firs case confirmation and the second the 40 day lockdown mark. Panel A and B: The vertical black dashed line corresponds to the beginning of the lockdown (March 16, 2020) and the the grey shading to the lockdown period, the solid blue line corresponds to the LOESS smoother and the blue shading corresponds to a 0.6 smoothing span around the LOESS smoother line.

## Discussion

In this nationwide time series analysis, we found that lockdown implementation was associated with a sudden reduction in all major forms of violent and accidental deaths (homicides, suicides and traffic accidents represented 79% of total violent and accidental deaths in 2019^39^) with a detected change in the post-lockdown trend of suicides in men. Nonetheless, we do expect rates to return to their pre-pandemic levels at some future point. The biggest immediate absolute reduction was seen in traffic accidents and the smallest in suicides, but of course this also reflects the higher absolute burden of traffic accidents as a cause of death.

These results are consistent with both the theoretical rationale behind lockdown and various early reports in other parts of the world. The change in lifestyle and behaviors associated with limited outside activities and economic shutdown must certainly play a role in the mechanism behind the acute change in the rates of these types of deaths. Falls in mobility have a natural impact on road traffic accidents, since people staying at home are at no risk for these events. The decreases in suicide and homicide are less obvious. Suicide might be expected to increase from economic and social stress and the disruption of daily routine^40,41^.

Expectations about homicide are also not so clear. As lockdown measures began, conventional crimes began to slow down around the world. Studies that evaluated the short-term effects of lockdown on different types of crime reports in Los Angeles and Indianapolis in the USA found a marked decrease in the robbery, burglary, and aggravated assault after the stay-at-home measures took place^18,42^.

Most homicides in men in Latin-America and around the world are associated with crime^43,44^, and since lockdown, both murder and crime decreased in the region^45^. In Mexico, murder rates, which started at a historic high in 2020, dropped dramatically almost halfway from the national average of 81 per day to 54 after social distancing measures were put in place^46^ and a similar pattern has been seen in other countries in the region. Although there is an initial drop and sustained low crime rate while in lockdown, a greater change is expected once lifted due to economic uncertainty. In our time series analysis, we also found a marked dropped in homicides after lockdown. Although this aligns with most reports in the region, this decrease in homicides contrasts with what is happening in some cities in the USA where crime is down, but murder is up, without a clear explanation of this divergence ^47^. Now in the post-lockdown period of our time series there is already the first hint of a subtle increase in the rates of homicides.

Most homicides in men are associated with crime, however, most homicides in women are hate crimes or feminicides. In Peru, the first cause of homicide in women is intimate partner violence, with 1 of every 5 women having the partner as the perpetrator ^48^. Lockdown and isolation measures to prevent the spread of COVID-19 have created greater risks for women living in situations of domestic violence^23,24,49^. Although we found a reduction in women’s homicides overall, this does not exclude the increase in other forms of intimate partner violence, rather, it reflects the conditions in which femicides occur. According to a study carried out in autopsies of women victims of violence, the most frequent place where the body was found was in public places, rivers, or open fields (43.4%), compared to the home (22.9%)^50^. Lockdown restrictions may have imposed an additional barrier in the occurrence of these tragic acts as police and military were constantly watching the streets.

Mental health during lockdown has also been a constant concern^13,15,16,51,52^. Some initial reports show the increase in suicides rate during this pandemic as a consequence of lockdown, financial stress, uncertainty, and isolation^52,53^. Two possible projection scenarios based on an increase in unemployment in Canada following the COVID-19 pandemic resulted in a projected total of 11.6 to 13.6 excess suicides in 2020-2021 per 100 000^54^. Our results show that after lockdown the immediate rate of suicides declined, however, men presented a detectable increase in the slope of suicides in the post-lockdown period. One of the factors may be the shutdown of the economic activities and the financial stress during the post-lockdown period. Nearly 81% of men in Peru have a paid job compare to 64% women^55^, and during the pandemic, the male working population in the country capital decreased by 47.3% and the female working population by 48.1%^56^. These initial changes in suicide trends may give us an idea of what might come next as a result of these social changes.

Traffic accidents were the type of violent death that decreased the most and this observation is consistent with many of the specific restrictions adopted, including the general limit of transit as well as the imposition of strict curfews at night and on Sundays. Other countries have also seen a decline in emergency room visits for trauma injuries related to traffic accidents after the lockdown^28,57^.

An increasing rate towards the end of the lockdown is more apparent for traffic accidents than for other types of death. This mirrors quite closely the changes observed in mobility trends. Both, traffic accident deaths and the mobility change present as U-shaped trends during lockdown, demonstrating that the lockdown measures were not fully adopted at the beginning and that they eased gradually towards the end. Other forms of violent death have also shown a decline and even appear to be in decline after the lockdown, however, as economic activities resume, this rate may recover the baseline level as with the other types of death.

This report constitutes an initial analysis of the trends in violent deaths and as such, we recognize some limitations. Although we found that there was no major change in the occurrence of deaths coded as “unknown” cause, after lockdown, underreporting may be possible for other coding variables. This is a nationwide analysis and some differences by region may not be captured. Peru has tremendous diversity of lifestyle between coastal, mountain and jungle regions, and data come from large cities and small rural communities with radically different rates of events. Competing risk due to COVID-19 is also a possibility although most of the violent deaths occur in the younger population, and not necessarily in the population at risk of dying because of COVID-19. For suicide, however, the populations may overlap to a greater extent^58^. Our analysis only considers the beginning and end of the lockdown thus the sudden initial drop that we have described may be accompanied by a sudden increase after the measures are lifted and a later follow-up analysis would be informative.

There is an urgency to consider and understand the myriad indirect mortality consequences of the policies adopted to respond to COVID-19. It is expected that some time after the lockdowns are completely lifted around the world, the lives lost from the impacts of these various policies on the economy, lifestyle, and mental health will outweigh the number of lives lost directly from infection. Indicators of this broad impact, including the types of violent and accidental deaths studied here, will be crucial for future decision-making^59^.

## Conclusions

Lockdown due to COVID-19 has impacted the rates of violent and accidental deaths, showing a sudden drop after its implementation and an increase in the slope of suicide in men in the post-lockdown period. The biggest change was seen in deaths as a consequence of traffic accidents. This initial drop should not be considered as encouraging, since just as there was a marked drop at the beginning, it is likely to be an equally sharp increase after the lockdown is lifted and the economic activities are resumed. The patterns for suicide and homicide are less intuitive, however, and reveal important clues about the causes and characteristics of these events. Policies should take into consideration other aspects of health that might be overlooked during this pandemic.

## Data Availability

Based on data from the Peruvian National Death Information System

https://github.com/renzocalderon/COVID_External_Deaths_Peru

